# The Role of Modelling and Analytics in South African COVID-19 Planning and Budgeting

**DOI:** 10.1101/2022.08.23.22279123

**Authors:** Gesine Meyer-Rath, Rachel A Hounsell, Juliet RC Pulliam, Lise Jamieson, Brooke E Nichols, Harry Moultrie, Sheetal P Silal

## Abstract

**Background:** The South African COVID-19 Modelling Consortium (SACMC) was established in late March 2020 to support planning and budgeting for COVID-19 related healthcare in South Africa. We developed several tools in response to the needs of decision makers in the different stages of the epidemic, allowing the South African government to plan several months ahead of time.

**Methods:** Our tools included epidemic projection models, several cost and budget impact models, and online dashboards to help government and the public visualise our projections, track case development and forecast hospital admissions. Information on new variants, including Delta and Omicron, were incorporated in real time to allow the shifting of scarce resources when necessary.

**Results:** Given the rapidly changing nature of the outbreak globally and in South Africa, the model projections were updated regularly. The updates reflected 1) the changing policy priorities over the course of the epidemic; 2) the availability of new data from South African data systems; and 3) the evolving response to COVID-19 in South Africa such as changes in lockdown levels and ensuing mobility and contact rates, testing and contact tracing strategies, and hospitalisation criteria. Insights into population behaviour required updates by incorporating notions of behavioural heterogeneity and behavioural responses to observed changes in mortality. We incorporated these aspects into developing scenarios for the third wave and developed additional methodology that allowed us to forecast required inpatient capacity. Finally, real-time analyses of the most important characteristics of the Omicron variant first identified in South Africa in November 2021 allowed us to advise policymakers early in the fourth wave that a relatively lower admission rate was likely.

**Conclusion:** The SACMC’s models, developed rapidly in an emergency setting and regularly updated with local data, supported national and provincial government to plan several months ahead of time, expand hospital capacity when needed, allocate budgets, and procure additional resources where possible. Across four waves of COVID-19 cases, the SACMC continued to serve the planning needs of the government, tracking waves and supporting the national vaccine rollout.

## INTRODUCTION

To date, South Africa has experienced four waves of COVID-19, with an official tally of more than 3,711,000 cases and 101,000 reported deaths as of May 2022 [1]. The country reported its first imported COVID-19 case on 5 March 2020, with subsequent rapid spread into all districts in the country. In response, the South African government implemented a five-level COVID-19 alert system, beginning with a full lockdown (Level 5) from late March 2020 [2]. The alert levels determine the extent of restrictions to be applied during the national state of disaster, which was initiated in March 2020 and ended two years later, in April 2022 [2]. The risk-adjusted approach was guided by several criteria, including numbers of infections and rate of transmission, health facility capacity, the extent of the implementation of public health and social measures (PHSM), as well as the economic and social impact of continued restrictions [2].

The South African COVID-19 Modelling Consortium (SACMC) was established at the end of March 2020 in response to a request by the South African National Department of Health (NDOH) to project the spread of the disease to support policy and planning in South Africa over the course of the epidemic. The consortium developed two models to project incidence, deaths, need for hospital beds at different levels of care, and the corresponding resources required. The models incorporated available data on COVID-19 cases, severity, and mortality, and served to inform the public and a range of decision makers, including the Ministerial Advisory Committee on COVID-19 (MAC) advising the Minister of Health, staff in the NDOH and National Treasury, officials in the provincial departments of health, and the private health sector.

This paper describes the 2-year process of continuously updating the models while collaborating with the diverse partners, and gives an overview of the model results and the consortium’s recommendations at the different stages of the South African COVID-19 epidemic. The aim is to highlight the dynamic, multidisciplinary nature of policy-driven modelling of an emergency in a country with severely constrained resources. A full description of the models developed during wave 1 is provided in [3], and our analysis of COVID-19 related hospitalisations in [4].

## METHODS

### The South African COVID-19 Modelling Consortium

The SACMC is a group of researchers from academic, non-profit, and government institutions across South Africa. Established in late March 2020 by the NDOH, its mandate is to provide, assess, and validate model projections to be used for planning purposes by the Government of South Africa. The SACMC’s work is coordinated by the South African National Institute for Communicable Diseases (NICD), which maintains the datasets used by the SACMC’s models. In addition to its core group of experienced infectious disease modellers and health economists, the SACMC convenes experts across a range of disciplines to provide insights, guide the selection of appropriate parameter values, ensure a close alignment to current clinical practice, and sense-check model outputs.

### Policy-driven modelling

Since its establishment in March 2020, the SACMC has provided policy-driven modelling and analytics support in response to the evolving priorities of decision makers across the different stages of the epidemic. Several tools were developed and adapted over time to meet these needs. Taken together, these tools supported the South African government at national and provincial levels to conduct timely resource planning, shift scarce resources, and implement appropriate PHSM.

At the start of the epidemic, the most pressing need was for short- and long-term projections of COVID-19 cases, including the number of severe and critical cases requiring hospital admission, and deaths under different PHSM scenarios. To fulfil these needs, the SACMC developed the National COVID-19 Epi Model (NCEM), a compartmental transmission model following a generalised Susceptible-Exposed-Infectious-Removed structure that accounts for disease severity (asymptomatic, mild, severe, and critical cases) and treatment pathways (outpatient, inpatient non-ICU and ICU care) [3].

The National COVID-19 Cost Model (NCCM), a companion model, used epidemiological outputs from the NCEM on the number of mild, severe and critical cases to project total COVID-19 resource needs and the associated impact on the national and provincial health budgets by incorporating information on the need for inpatient and outpatient resources (including their baseline availability and how to scale availability with the size of the epidemic). Resource projections covered drugs, diagnostics, ventilators, oxygen supply, field hospitals and other hospital infrastructure, staffing requirements, and additional mortuary space; model extensions included the quantification and cost of vaccines under different vaccination scenarios. These projections informed the development of resource quantifications and budgets, allowing timely negotiation with manufacturers and preparation of contracts for the additional resources anticipated based on precise quantifications, with the exact volumes required being regularly updated based on the latest model outputs.

As the epidemic progressed, in addition to the ongoing projections described above, priority was placed on resurgence monitoring, estimating the impact of relevant emerging variants of concern (Beta, B.1.351, which first emerged in South Africa; Delta, B.1.617.2; and Omicron, B.1.1.529, which was again first identified in South Africa), and modelling to inform the government’s vaccination procurement and rollout strategy. Figure 1 shows the different tools developed over the course of the epidemic in South Africa, which are discussed in further detail below, and the timing of their main outputs.

**Fig 1.**
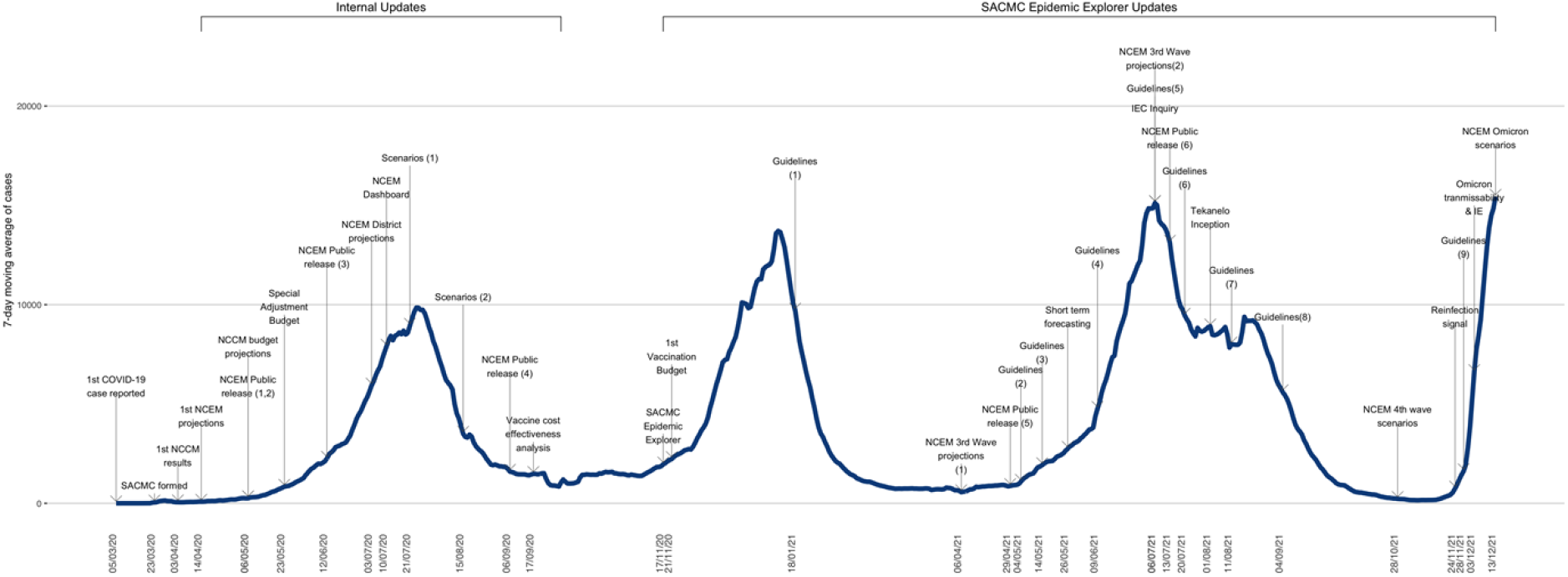
Select SACMC modelling and analysis contributions: Timeline from March 2020 to December 2021. See Table S1 for additional information.

### Model users

The SACMC’s modelling outputs have been used by a range of strategic and operational decision makers (Table 1). Different departments within the NDOH used NCEM outputs for purposes ranging from the quantification of drug volumes required for inpatient and outpatient care by the Affordable Medicines Directorate to the estimation of additional mortuary and graveyard spaces by the Environmental Health Directorate. Private sector initiatives such as the National Ventilator Project, which sourced ventilation equipment for public and private hospitals in South Africa, used our model outputs as well. Analysts coordinated by the Reserve Bank, the South African central bank, used SACMC model outputs to predict the macro-economic impact of the epidemic under different scenarios.

**Table 1:** Uses of NECM and NCCM outputs.

| User | Purpose |
| --- | --- |
| NDOH: |  |
| Ministerial Advisory Committee | Policy advice |
| Facility Readiness Committee | Number of beds and expansion of facilities |
| Human Resources for Health team | Staff required per level |
| National Ventilator Project | Number of ventilators needed |
| Oxygen planning team | Oxygen required |
| Affordable Medicines Directorate | Drug quantities |
| National Health Laboratory Service | Number of tests and testing priorities |
| Environmental Health Directorate | Number of mortuary containers |
| Provincial DOHs | Local planning and resource quantification |
| National Treasury | COVID-19 health budgets |

The resurgence monitoring tool was used by planners and technical advisory teams to declare a resurgence of cases and respond according to the guidelines detailed in the NDOH Resurgence Plan [5], while the third and fourth wave scenario modelling and short-term forecasts were used primarily to estimate hospital admissions ahead of a third wave driven by the Delta variant in June 2021, and a fourth wave driven by the Omicron variant in November 2021.

Outputs of the NCCM and vaccination cost models were used by the National Treasury to make decisions regarding the additional budget allocation required for COVID-19. Model outputs in part informed a budget allocation of 1.42 billion USD for COVID-19 specific health care announced by the South African president at the end of April 2020 [6], most of which was financed through the reallocation of the existing health budget [7]. Additionally, a group of experienced public finance specialists was trained to work with provincial Departments of Health to update the model with provincially specific data, including the baseline availability of resources, prices, and need for resources that were independent of the course of the epidemic, such as personal protective equipment and isolation and quarantine facilities.

### Key parameters and data sources

The parameter values for the early versions of the NCEM were based on literature and data from other countries as well as local expert opinion regarding the types, duration, and outcomes of hospital treatment. Parameter values and assumptions were regularly updated as the scientific knowledge base on COVID-19 expanded. As South African data became available, parameters were adjusted to reflect the local context. Parameter selection was guided by ongoing input from clinicians, virologists, intensive care specialists, immunologists, and epidemiologists on the SACMC.

Table 2 juxtaposes the initial set of parameter values from late April 2020 and the updated set from September 2020; Table 3 gives an overview of the main data sources.

**Table 2:** Key NCEM parameter values and their evolution between April and September 2020.

| Parameter | April 2020 |  | September 2020 |  |
| --- | --- | --- | --- | --- |
|  | Value* (range) | Sources | Value* (range) | Sources |
| **Infection severity and transmission\*\*** |  |  |  |  |
| Proportion of cases that are asymptomatic | 75% | [9-11] | 75% (70% - 80%) | [8-11] |
| Relative infectiousness of asymptomatic cases | - | - | 80% (77.5%, 82.5%) | [12-14]<br>Estimated through calibration to admissions and fatalities count data (DATCOV) [15] |
| Mild to moderate cases among the symptomatic | (95.64%, 96.78%) | adjusted based on [17] | (94.55% - 97.13%) | Estimated through calibration to admissions and fatalities count data (DATCOV) [15] [16] |
| Severe cases among the symptomatic | (2.46%-3.64%) |  | (2.58% - 5.00%) |  |
| Critical cases among the symptomatic | (1.16%-1.45%) |  | (0.18% - 0.55%) |  |
| Proportion of cases that are fatal | (0.30%, 0.412%) | [17], [18] |  |  |
| **Timeframes & treatment duration** |  |  |  |  |
| Time from infection to onset of infectiousness | 4 days (2-9) | [18], [19-23]<sup>1</sup> | 2 days (1-3) | [24, 25-34] with input from the National COVID-19 Modelling Consortium |
| Time from onset of infectiousness to onset of symptoms | 2 days (1-4) |  | 4 days (3-5) |  |
| Duration of infectiousness from onset of symptoms | 5 days |  | 5 days (4-6) | [35,36] |
| Time from onset of mild symptoms to testing | 4 days (2-4) |  | 4 days (3-5) | [25, 26, 18-21, 23] |
| Time from onset of symptoms to hospitalisation | 5 days (4-8) |  | 5 days (4-6) |  |
| Time from onset of symptoms to ICU admission | 9 days (8-17) |  | see below |  |
| Duration of hospital stay | 12 days (7-16) |  |  |  |
| Duration from ICU admission to discharge | 18 days (14-18) |  |  |  |
| Duration from ICU admission to death | 5 days (4-7) |  |  |  |
| Time in non-ICU (never ICU) to death/recovery |  |  | 8 days (4-12) | Lengths of stay: values and ranges sourced from NICD |
| Time in non-ICU for those destined for ICU |  |  | 0 days (0-2) |  |

<sup>1</sup> with additional input from analysis of NICD data

|  |  |  |  |  |
| --- | --- | --- | --- | --- |
| Time in ICU for those ventilated and destined to die |  |  | 14 days (7-27) | COVID-19 Hospital Sentinel Surveillance database (DATCOV) [15] [4] |
| Time in ICU for those never ventilated and destined to die |  |  | 11 days (7-18) |  |
| Time in ICU for those ventilated and recovered |  |  | 19 days (15-37) |  |
| Time in ICU for those never ventilated and recovered |  |  | 5 days (1-10) |  |
| Time in non-ICU for those who were in ICU and recovered |  |  | 0 days (0-6) |  |

**Table 3:** Summary of NCEM data sources.

|  |
| --- |
| National case and hospitalisation data from the South African National Institute for Communicable Diseases |
| Statistics South Africa projected 2020 district population projections [37] |
| Coronavirus COVID-19 (2019-nCoV) Data Repository for South Africa, Data Science for Social Impact Research Group @ University of Pretoria [38] |
| Vodacom Mobile Event Database |
| Google COVID-19 Community Mobility Reports |
| Published and pre-print academic literature (cited in Table 2) |
| Expert input from members of the SA COVID-19 Modelling Consortium, and https://sacoronavirus.co.za/category/press-releases-and-notices/ |

The NCCM used three types of input data: 1) The type and required quantities of resources, such as the number of inpatient beds projected by the NCEM, human resources at all care levels, oxygen, oxygen delivery devices, SARS CoV-2 tests, infection control and prevention infrastructure; 2) the public-sector prices of these resources; and 3) the baseline volume of resources available for the COVID-19 health response for all items where the quantities required exceeded existing resource levels, such as in the case of hospital beds or ventilators. Costs were evaluated from the provider perspective, in this case the South African government. Despite the need to provide COVID-19 testing and care in both the public and the private sectors, this perspective is appropriate given that contracting arrangements were put in place to ensure private-sector services were offered at charges similar to public-sector prices. As a result, we used public-sector prices and salaries throughout, based on the most recent tariffs and public tenders (Table 4).

**Table 4:** Summary of NCCM data sources.

| Intervention/ ingredient | Type of target population | Data sources |  |  |
| --- | --- | --- | --- | --- |
|  |  | Target population | Cost | Baseline capacity |
| PPE | Healthcare workers at all levels | DHIS | Own analysis | NDOH |
| ICU beds, staff and linen | Critically ill cases | NCEM | Own analysis | NDOH |
| Ventilators | Critically ill cases (subset) | NCEM | Public tender | NDOH |
| Testing | All cases who present for testing | Own analysis | NHLS | NHLS |
| Community health workers (CHW) | 10,000 | DHIS | Public salaries | - |
| CHW supplies | Thermometers for 10,000 CHW | - | Public tender | - |
| Isolation |  | - | Own analysis | - |
| O2 | Severely + critically ill cases | NCEM | Own analysis | NDOH |
| Hospital beds, staff and linen | Severely ill cases | NCEM | Own analysis | - |
| Drugs | All cases (by level of severity) | NCEM | Own analysis | - |
| PHC staff | 70% of symptomatic mild cases | NCEM | Own analysis | - |
| Fever clinics | NDOH | NDOH | NDOH | - |
| 30-bed wards | NDOH | NDOH | NDOH | - |
| Mortuary cupboards and cabinets | Deaths by municipality | NCEM | NDOH | NDOH |

**Table 5.** The projected COVID-19 health budget for financial year 2020/21.

| Budget item | Description | Scenario with 20,000 additional ICU beds |  |
| --- | --- | --- | --- |
|  |  | Total cost [millions 2020 USD] | % of total cost |
| PPE | Personal protective equipment for healthcare workers at all levels | 360 | 18% |
| Testing | PCR tests only; no new laboratory instruments or extra staff | 145 | 7% |
| Central functions | Port Health and surveillance | 23 | 1% |
| Intensive Care Unit beds | incl. additional beds required, linen and staff costs | 426 | 21% |
| Ventilators | Additional ventilation equipment required for inpatient care | 74 | 4% |
| Oxygen | Oxygen cylinders for inpatient care, excludes oxygen equipment | 179 | 9% |
|  |  | Total cost [millions 2020 USD] | % of total cost |
| Hospital beds | incl. additional beds required, linen and staff costs | 9 | 0% |
| Drugs | at ICU, general wards and primary healthcare clinics | 211 | 10% |
| PHC staff | for screening, testing, clinical assessment, post-test follow-up | 32 | 2% |
| CHW supplies | 1.1 thermometers per community healthcare worker | 7 | 0.3% |
| Isolation facilities | Upgrading and repurposing of hotel and conference facilities to be used as facilities for isolation of mild cases | 60 | 3% |
| Fever clinics | 1000 units to be added to primary health centres and community health centres | 218 | 11% |
| 30-bed COVID-19 wards | Attached to existing hospital or to field hospital | 299 | 15% |
| **TOTAL** |  | **2,043<sup>2</sup>** |  |

### Evolution of modelling purpose, structures, and tools across the epidemic

#### Projections and scenarios for short- and long-term COVID-19 burden

Government set three priorities during the first wave of the COVID-19 epidemic in South Africa: 1) to generate short- and long-term projections of COVID-19 cases, estimating the pace at which cases might increase and spread between provinces; 2) to project the expected number of severe and critical cases leading to hospital admission, as well as estimates of the corresponding resource requirements; and 3) to compute the cost of the health sector response to the epidemic at a provincial and national level in order to inform the adjustment of the health budget and the flow of resources around the country.

In order to fulfil the first and second priority, we developed the National COVID-19 Epi Model, a compartmental transmission model to estimate the total and reported incidence of COVID-19 in the nine provinces (and later, 52 districts) of South Africa. It was designed to simulate the impact of different behavioural scenarios, inform resource requirements, and predict where gaps could arise based on the available resources within the South African health system. The model follows a generalised Susceptible-Exposed-Infectious-Removed (SEIR) structure accounting for disease severity (asymptomatic, mild, severe, and critical cases) and the treatment pathway as shown in **Error! Reference source not found**..

**Fig 1:**
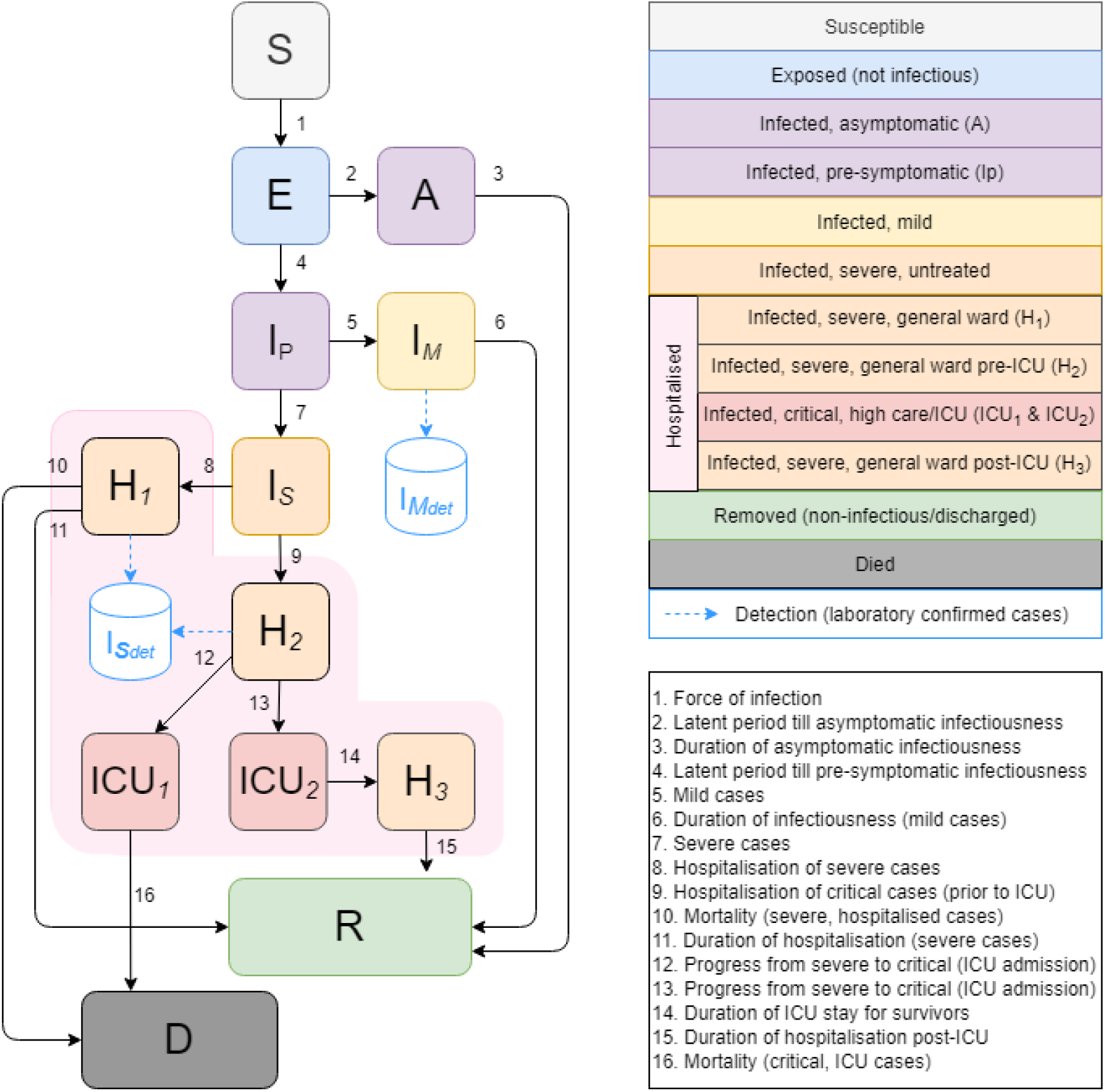
Original NCEM model structure (v1) Version 1 of the NCEM v1, stratified at the provincial level, accounted for the clinical profile of SARS-CoV-2 and the hospital care pathway (Figure S1). With limitations on general ward and ICU capacity, the model was extended in v2 to account for limited access to hospital level care and restricted capacity in wards at a provincial level (Figure S2). With a need for planning support at a finer spatial granularity and lifting of restrictions on intra-country travel, a stochastic version of the NCEM (v3) was extended to cover the 52 districts of South Africa, linked through a connectivity matrix formulated by mobile data over time. NCEM v4 was necessary after the detection of the Beta variant in South Africa (Figure S3). The model was further stratified to include 7 age groups across three subpopulations of interest: healthcare workers, the population with comorbidities and everyone else. Vaccination was included to account for vaccine effectiveness against infection and severity for a generic vaccine. In anticipation of another variant and with the vaccination programme being implemented for healthcare workers NCEM v5 was developed to include a third variant stratification for this hypothetical new variant, which was updated at the beginning of the third wave to represent early data on the characteristics of the Delta variant (Figure S4). With NCEM v6, vaccination was included in greater detail with specific vaccination types in mind, differentiating between vaccine effectiveness in those with prior infection vs the immunologically naïve (Figure S5). Lastly, just before the fourth wave the NCEM was once again updated to v7 with a fourth stratification for another hypothetical new variant, producing various scenarios driven by different joint assumptions regarding its transmissibility and immune escape properties (Figure S6). This version was updated to the characteristics of the Omicron variant within days of Omicron being named.

The NCEM was originally developed as a deterministic compartmental model, where the characterisation of uncertainty changed over the course of the pandemic. Early on in the first wave, when uncertainty in parameter values was large and the least complex framework of the NCEM was implemented, uncertainty in parameter values was explored through sensitivity analyses and uncertainty bounds estimated through random draws from specified parameter distributions. When the need arose to expand the model to a finer spatial granularity in v3, the NCEM was additionally implemented stochastically to account for greater uncertainty at small geographical levels. The inclusion of variants and vaccines into NCEM versions 4 onwards brought an additional layer of uncertainty in projection modelling, and scenario analyses were employed to depict uncertainty in the outcomes of unknown vaccine and variant characteristics.

#### Resource planning

The National COVID-19 Cost Model (NCCM) takes inputs from the NCEM and cost inputs based on data from existing sources that were adapted to represent the type, number and prices of ingredients required in South Africa’s COVID-19 response. It calculates annual budgets for the NDOH’s response to COVID-19, allocating costs at the level of the provinces as well as NDOH, incremental to existing resources such as hospital beds and staff contingents.

The NCCM was updated with new NCEM results whenever they became available and was changed to incorporate additional interventions when deemed relevant by policy makers and planners (for example, temporary inpatient infrastructure such as field hospitals and add-on clinic space). Additional updates included new clinical interventions once they were incorporated into national COVID-19 management guidelines (such as dexamethasone treatment and high-flow nasal cannulae treatment), and prices and quantities as new tenders and data on actual resource use became available. For example, in July 2020 we adjusted our assumptions regarding inpatient length of stay downwards from the initial estimates based on international literature to results from our analysis the South African hospital sentinel surveillance database [15] (for more detail regarding the methods see [4]); and inpatient costs were updated based on a more detailed analysis based on South African ingredients and prices [41].

#### Dashboard for disseminating model updates

Between early April and early September 2020, the NCEM was updated frequently, and results made available to stakeholders within the South African government. Reports on a subset of these updates were additionally made public. To aid the timely dissemination of projections, we developed an web-based interactive application, the National COVID-19 Epi Model Dashboard (https://masha-app.shinyapps.io/NCEMDashboard), which visualised the most important NCEM outputs on projected cases, hospitalisations and deaths. These included active cases and cumulative detected cases for symptomatic, severe, and critical cases; hospital beds needed and cumulative admissions for non-ICU, ICU ventilated, and ICU non-ventilated; and cumulative deaths.

#### Resurgence monitoring

At the beginning of the second wave of COVID-19 cases in September 2020, driven by the Beta variant which originated in South Africa, resurgence monitoring became a priority. The SACMC assisted in the development of a set of resurgence metrics to support planners and technical advisory teams to declare a resurgence or wave and respond according to the guidelines detailed in the Ministry of Health’s resurgence action plan [5]. To facilitate effective communication and dissemination of these metrics, we developed a second web-based dashboard called the SACMC Epidemic Explorer in two versions: a sub-district level version to guide planners at all government levels, and a district-level version (www.SACMCEpidemicExplorer.co.za) to inform the public. It allows users to explore the COVID-19 epidemic in South Africa, analyses resurgence risk, presents metrics such as estimates of case levels, percentage change in cases and periods of consistent growth to prepare for future outbreaks, and monitor confirmed COVID-19 hospital admissions and deaths. We measured the metrics’ performance against three main criteria: whether the resurgence metrics were consistent in their messaging; whether the metrics provided sufficient early warning; and whether the metrics relaxed the severity of their messaging at a suitable time and pace after the peak of the epidemic.

#### Short-term forecasting

Towards the end of the second wave, short term forecasts of COVID-19 cases, hospital admissions, and of the likelihood of when provinces would meet the third, then fourth wave criterion were developed in order to improve situational awareness and inform government resource planning. These two-week case and admission forecasts were updated weekly and three times weekly, respectively, and displayed on the SACMC Epidemic Explorer for easy access.

## RESULTS

### Findings of the National COVID-19 Epi Model (First wave)

In May 2020, using version 1, the model projected an estimated 8.01 and 8.62 million laboratory-confirmed cases, and 40,223 and 43,759 deaths, in the optimistic and pessimistic scenarios, respectively, by 1 October. Active cases were estimated to peak in early July in the pessimistic, and mid-July in the optimistic scenario, with a maximum number of between 72,281 and 77,899 hospital beds, and 31,656 and 24,150 ICU beds in use at peak. Total cumulative incidence was estimated to reach between 48.7 and 51.7 million cases (symptomatic or asymptomatic) by 1 October (i.e., an attack rate of 82-88%), with between 8.01 and 8.62 million detected cases, assuming a detection factor of 1 in 6 cases. While these projections incorporated a certain level of effectiveness of PHSM in their scenarios, they underestimated the severity and effect of government restrictions and population adherence to them and thus assumed that all exposure would happen during a single wave. A full set of projections for all nine provinces is available in our detailed reports [42, 43].

The updates in version 2 that took into account the variation in timing and level of peaks of the epidemics between the provinces and between the districts in each province resulted in a national peak in cases at a a similar time (i.e., mid-August 2020) to the optimistic scenario from version 1, but at a lower level. While the model projected a concomitant lower peak in the need for hospital (non-ICU) and ICU beds at a national level, bed capacity was still expected to be breached or overwhelmed in all provinces. We noted that increasing capacity to accommodate patients in hospital could allow the country to better leverage new therapeutic options such as high-flow oxygen and dexamethasone, which had the potential to improve mortality outcomes.

In version 3, we modelled the impact of four behavioural scenarios on the four provinces with the most advanced epidemics: Western Cape, Eastern Cape, Gauteng and KwaZulu-Natal. When applying each scenario, we noticed that each in turn led to either earlier and/or lower peaking of cases than our original projections, with the exception of the scenario in which the behavioural response threshold was assumed to be 110 deaths per day, which peaked at roughly the same level but shifted the peak forward slightly in all three provinces. The full analysis detailing the behavioural scenarios is available in [44].

The updated version 4 from early September 2020 estimated that there had been 15.2 million infections by September, equating to 25.5% (uncertainty range: 22.0%-28.6%) of the population- a much lower first-wave attack rate than estimated in our first version. Under the moderate testing scenario, cumulative detected cases were estimated to continue to grow until between 570,000 and 1.2 million cases by early November (and only marginally so thereafter), depending on testing rates. The peak number of general hospital (i.e., non-ICU) beds in use was estimated to have been reached in early-August, at around 8,000 beds (when around 12,500 beds would have been needed). The peak number of ICU beds in use was estimated to have been reached around the same time, with around 1,100 beds-although more than 2,000 beds would have been needed. Total deaths are estimated to continue to increase until early November when the cumulative number of all deaths would reach 37,000 (of which 16,000 would have been in hospital); thereafter the growth rate was estimated to be very low [44].

### Findings of the National COVID-19 Cost Model (First wave)

Given the regular updates to the NCCM’s structure and inputs, and the ongoing changes to the country’s COVID-19 management policies and clinical guidelines, model results changed almost weekly for the duration of the first wave. Amongst the many updates, we report on a version in the end of May 2020, which used input from the first NCEM version to inform an additional allocation to COVID-19 related healthcare for financial year 2020/21.

The NCCM estimated that the budget required for the COVID-19 health response for financial year 2020/21 would be around 2.1 and 2.7 billion USD under the NCEM’s optimistic and pessimistic scenario, respectively (**Error! Reference source not found**.). Scenarios differed in the cost of those budget items that were directly linked to the number of projected cases, in particular inpatient care (ICU and non-ICU beds, ventilators, and oxygen) and drugs at all levels of the healthcare system. In both scenarios, the largest contributor to total cost were the procurement and staffing of additional ICU beds (21% and 26%, resp., in the optimistic and pessimistic scenario), personal protective equipment (PPE) for healthcare workers at all levels (18% and 14%), and prefabricated facility infrastructure discussed at that time, such as additions to primary healthcare clinics for the management of non-severe patients (“fever clinics”) (11% and 9%) and 30-bed stand-alone COVID-19 wards as additions to hospitals (15% and 12%).

Based on these results, about 1.5 billion USD was added under the COVID-19 Special Adjustment Budget in June 2020, funded through a combination of reprioritisation of funds from other departments and within the provincial health budgets, and increased lending [7]. Most of this budget was used in expenditure on inpatient care and PPE, although over 12 instead of the originally estimated 6 months, and allocated slightly differently: Few of the large infrastructure projects were implemented, except for the construction of three large field hospitals in the worst-hit provinces, in two cases through the repurposing of existing buildings, and with completion so delayed that very few patients were admitted. In total fewer ICU beds were added due to severe constraints in the trained staff needed to staff them.

### Findings from resurgence modelling, forecasts and scenario modelling (Wave 2 onwards)

The resurgence monitoring dashboard provided continuous updates three times a week on the state and growth of the epidemic across provinces, districts, and sub-districts from December 2020 onwards. The metrics on the dashboard classified all areas as being in a state of control, alert or response, which triggered a series of actions laid out in a Department of Health workplan [5]. The short-term forecasts provided two-week forecasts on the trajectory of cases and admissions at the provincial level from May 2021 on, and placed these projections within the context of the relationship between weekly admissions and hospital-based case fatality rates that was seen in prior waves, with one notable exception: At the beginning of the fourth, Omicron, wave, we did not provide admissions forecasts, due to uncertainty regarding the relationship between cases and admissions under this novel variant.

The scenario modelling for the third wave which accounted for province-level data on seroprevalence and the dominance of new variants estimated that across most provinces and behavioural scenarios, the peak admissions of the third wave would be lower than that of the second wave. We however saw that a slow, weak behavioural response would increase admissions for severe or critical COVID-19 cases across most age groups. Younger age groups were expected to have fewer admissions than in the second wave in most scenarios. Our analysis by province showed substantial variation of the size of the third wave between provinces, reflective of different age distributions, seroprevalence and prevalence of comorbidities, with the third wave being highest in more urban provinces, due to the higher concentration of working-age adults and people with co-morbidities. Across provinces, the time from an initial increase in transmission to the peak was estimated to be on average 2-3 months. A full report detailing scenario projections for each of the nine provinces is available in [45, 46].

Lastly, our scenarios for the fourth wave projected that, across a large range of assumptions regarding immune escape properties and transmissibility a hypothetical new variant, loss of protection against severe infection, and increase in contacts, hospital admissions during the 4th wave would be less than during the 3rd wave, given high population levels of combined immunity from, in particular, previous infection and, to a lesser extent, vaccination (owing to low vaccine coverage at the time). This remained true in our updated sets of scenarios incorporating the higher transmissibility and immune evasion properties of Omicron compared to previous variants.

## DISCUSSION

The process of providing policy-relevant research on the evolution and impact of an aggressive novel pathogen required a relevant, timely and responsive modelling approach; ongoing re-assessment of the availability of local and international data and evidence; and effective, transparent navigation of uncertainty. Through our experience providing modelling and analytics support throughout the COVID-19 epidemic in South Africa, we learned the following lessons:

### The modelling approach needs to be relevant and responsive to evolving policy priorities

Over the course of the epidemic, the type of questions that are most important to decision makers changes. Modellers need to work closely with model users so that the outputs developed are relevant and timely, serving the dynamic needs of the user. Further, establishing strong relationships and embedding continuous engagement with stakeholders throughout the modelling process is fundamental to accelerating the incorporation of modelling output into decision making. In South Africa, policy needs changed from long term projections of the shape and peak of the first wave, to modelling of the impact of behaviour to explain diversions from expectation, to real-time resurgence monitoring during the ensuing waves (each driven by a new variant), incorporating all previous learnings into third wave projections (alongside two-week forecasts of hospital-relevant data), providing scenarios that included a potential novel variant with immune escape properties just ahead of the fourth wave driven by Omicron, and, finally, updating our fourth wave scenarios with the results of our own rapid-fire analysis of Omicron properties within days of the emergence of this variant. The approach discussed in this paper demonstrates the value of the adaptive nature of our policy-driven modelling work.

### Modellers need to be flexible in the tools they apply

At different stages of the epidemic, the varying availability of data and most pressing policy questions should inform the selection of an appropriate modelling or analytics approach. Short-term modelling can account for rapid and frequent changes, quickly incorporating new data, and enabling short lead time between updates and communication of findings. This is particularly relevant for important but rapidly changing inputs such as available hospital capacity, which has been difficult to estimate throughout the epidemic as facility capacity changed regularly. [42-44]. Additionally, we realised that, while our initial projections had intentionally projected the number of beds *needed* without taking capacity constraints into account, as well as the cost of providing oxygen and other resources to all critical cases, the actual capacity to accommodate severe and critical cases was much lower, leading to the addition of outputs (including budgets) based on actual bed *use*. The ability to project both of these during the first wave, in addition to predicting the extent of the health burden across provinces during the third and fourth waves, and monitor resurgence patterns and bed occupancy at the sub-district level, were possibly amongst our most useful contributions. However, even these outputs’ usefulness was limited by the non-fungibility of healthcare resources in many instances: The strongest constraint on inpatient bed availability was human resources, which could not be easily shifted and much less created over a short time span. And more than two thirds of deaths happened outside of hospitals, potentially pointing to COVID-19 patients being disheartened by reports of locally overwhelmed hospitals [47-49], being turned away by overburdened emergency rooms, becoming too sick too quickly to seek care in time, or dying in transit where emergency transport was scarce.

We also learned that it is in the hands of the modeller to judge if sufficient data are available and/or modelling can responsibly support the decisions to be made. Ahead of the second wave in South Africa in October 2020, model predictions of the shape and timing of the peak of this second wave were urgently requested. The SACMC decided to first assess the driving forces behind the resurgence, considering factors such as increased mobility, PHSM fatigue, lower seroprevalence or new variants. In the absence of additional information at the time (with the Beta variant only discovered in December 2020), the SACMC made the difficult decision not to produce model-based projections. Instead, we choose to develop a set of metrics that could detect and monitor the second wave. Further updates to the NCEM were only resumed in anticipation of the third wave and the vaccine roll-out programme, once more information was available. And again, at the beginning of the fourth wave, our use of epidemiological tools such as reinfection analysis [50] allowed us to quickly produce bounding estimates for Omicron’s combined transmissibility and immune escape properties [51], which in turn provided reliable estimates of bed needs for this wave.

### Models should take local context into account

There are several challenges to modelling in low- and middle-income countries in general. Data systems and surveillance infrastructure are often underdeveloped. Constrained resources, especially during health emergencies, lead to overwhelmed hospital staff who are unable to feed data systems in real time. This can cause delays and errors in reporting. Heterogeneity in population characteristics and access to health infrastructure are vital elements in the disease ecosystem and essential inputs to disease models. Infectious disease models should account for local context such as data availability, health systems dynamics, demography, contact patterns, acceptability of interventions, and cultural influences. In countries where data availability is scarce, it is important to understand what adaptations are needed and how models may perform under different data constraints, and to communicate the resulting uncertainty effectively. As such, the adaptation of plug-and-play models from resource-richer countries remains an inferior option, as these are usually ill-equipped to provide reliable, ongoing, real-time decision-making support tailored to local needs, data and epidemic specifics. One example of this is our mis-specification of mortality rates in v1 of our model which was a result of a misinterpretation of international data on case fatality rates-in March/ April, 2020, the only available data [42]. Incidentally, updating the model with locally derived mortality rates after the first South African wave resulted in very similar, albeit more correct, estimates of the number of overall deaths [44]. Another is our decision to not incorporate the impact of individual PHSM, despite clear and regular requests from policy makers to help with decisions regarding which individual restrictions would still be necessary. This decision was taken because, despite regular reviews, we did not find a robust enough dataset applicable to a low- or middle-income setting or South Africa specifically regarding the impact of these measures.

### Communicating uncertainty clearly and transparently is vital when reporting model findings

Managing expectations regarding the limitations of models, the quality of data and the assumptions used improves the likelihood of model outputs being used responsibly and appropriately by decision makers. We used several widely accepted approaches for doing this, including using scenario and sensitivity analyses, and clearly marked uncertainty ranges for input parameters and results. We found that uncertainty, while central to our understanding of our role as modellers, was not always useful to our audience of health planners; especially where additional resources had to be made available, users opted to use our median estimates in order to inform budgets and order exact quantities.

## CONCLUSION

In developing the NCEM, NCCM, and a number of dashboards and additional outputs such as reports and briefing materials, the SACMC supported national and provincial government to plan several months ahead of time, expanding hospital facilities where needed, and procuring additional resources. As the country is continuing its path towards endemicity, the SACMC continues to serve the planning needs of the government, tracking the development of cases and admissions and developing models to further support the national vaccine rollout.

Disease modelling was a source of regularly updated scientific evidence for decision making in the South African epidemic. Whilst much progress was made in developing models rapidly in an emergency setting, many challenges remain and need to be overcome to incorporate local context, needs of policymakers and sub-optimal data systems and build disease modelling capacity to better prepare for future health emergencies.

## Data Availability

All data are available in publicly accessible reports under https://sacmcepidemicexplorer.co.za/.

## ACKNOWLEDGEMENTS

The authors acknowledge the support of the National Department of Health and all planners, stakeholders and decision-makers who made use of these data systems and analyses to respond to the COVID-19 epidemic in South Africa. A large number of colleagues within the National Institute of Communicable Diseases, the Ministerial Advisory Committee on COVID-19, the Clinton Health Access Initiative, provincial and national Departments of Health, and National Treasury alerted us to policy-relevant questions, assisted the development of our models by providing data, discussing details of model structure, and reviewing assumptions and results. We are especially grateful to decision makers and planners at all levels of the health system who continuously used our outputs in the management of COVID-19.

